# A 56-Day Randomized, Double-Blinded, and Placebo-Controlled Clinical Assessment of Scalp Health and Hair Growth Parameters with a *Centella asiatica* Extracellular Vesicle and Growth Factor-Based Essence

**DOI:** 10.1101/2025.09.10.25335404

**Authors:** Tsong-Min Chang, Chung-Chin Wu, Huey-Chun Huang, Ji-Ying Lu, Ching-Hua Chuang, Pei-Lun Kao, Wei-Hsuan Tang, Wang-Ju Hsieh, Luke Tzu-Chi Liu, Wei-Yin Qiu, Ivona Percec, Charles Chen, Tsun-Yung Kuo

## Abstract

Hair loss and scalp dysfunction are widespread concerns with limited safe and effective long-term care options. This randomized controlled study evaluated the effects of a topical scalp care essence formulated with *Centella asiatica* extracellular vesicles (*C. asiatica* EV) and recombinant Fc-fusion long-acting insulin growth factor-1 (rIGF-1) and fibroblast growth factor-7 (rFGF-7) on scalp health and hair growth. Sixty healthy adult participants aged 18 to 60 years were randomly assigned into five groups: (1) Placebo control; (2) Base formula consisting of 0.1% caffeine and panthenol; (3) Base formula with long-acting rIGF-1 and rFGF-7; (4) Base formula with *C. asiatica* EV; and (5) Base formula combined with rIGF-1, rFGF-7, and *C. asiatica* EV. Participants applied the assigned product once daily for 56 days. Parameters, including sebum content, hair length, thickness, and density, and hair loss, were assessed at baseline and Days 14, 28, 42, and 56. Outcome measures included sebum content, hair density, scalp condition, hair length, and overall hair loss. The formulation with *C. asiatica* EV and growth factors was found to be most effective, with all parameters increased significantly on Day 56 compared to the placebo. This study highlights the potential of plant-derived EVs combined with growth factors as a synergistic strategy for promoting scalp and hair health.

## Introduction

Hair loss and hair thinning are globally prevalent issues. Statistics show that up to 50% of men and 30–40 % of women will experience some degree of androgenetic alopecia (AGA) during their lifetime [1]. This condition not only affects physical appearance but is also associated with significant psychological stress, anxiety, reduced self-confidence, and can even lead to a decreased quality of life and social difficulties [2, 3]. Among various forms of hair loss, androgenetic alopecia is the most common. Its pathogenesis involves a complex interplay of genetic predisposition, androgen regulation (such as the action of DHT and its receptor), and follicular microenvironment signaling pathways (including Wnt/β-catenin, TGF-β, IGF-1, etc.) [4, 5].

Although conventional treatments like minoxidil and finasteride are widely used and FDA-approved, their effectiveness requires long-term use. Minoxidil can prolong the anagen phase of hair follicles, but discontinuation often leads to rebound hair loss within a few months [6]. Finasteride may cause sexual dysfunction, hormone-related side effects, and poses a risk of teratogenicity if pregnant women are exposed [7, 8]. As a result, clinical concerns regarding safety, durability, and side effects have driven researchers to seek alternative therapies, including the use of growth factors (such as IGF-1 and FGF-7), stem cell-derived exosomes, and low-level laser therapy [9–12].

In recent years, advances in regenerative medicine and molecular biology have expanded hair follicle stimulation therapies to include growth factors, plant-derived bioactive compounds, and exosomes. Among these, insulin-like growth factor 1 (IGF-1) and fibroblast growth factor 7 (FGF-7; also known as keratinocyte growth factor, KGF) are considered two key bioactive factors. They have been shown to promote dermal papilla cell proliferation, prolong the anagen phase, and facilitate hair follicle regeneration [12–15]. IGF-1 is primarily synthesized by dermal papilla cells, hair matrix cells, and dermal fibroblasts [16]. It exerts proliferative and anti-apoptotic effects on follicular keratinocytes via the PI3K/AKT signaling pathway [13, 17, 18]. Additionally, IGF-1 is involved in regulating the differentiation of outer root sheath cells [13] and possesses antioxidant and anti-inflammatory properties [19, 20]. It prolongs the hair growth phase [14] and inhibits the transition of hair follicles into the catagen phase [13].

FGF-7 plays a role in epithelial–mesenchymal interactions and promotes the proliferation of hair follicle epithelial cells [12]. It also stimulates the growth of hair matrix cells and participates in the regulation of outer root sheath cell differentiation [15]. FGF-7 promotes hair growth by activating the MAPK/CREB signaling pathway and interacting with the Wnt/β-catenin signaling cascade [21, 22]. Like IGF-1, FGF-7 prolongs the anagen phase [12] and prevents the transition into the catagen phase [23]. Thus, FGF-7 has a critical role in regulating the hair cycle and promoting hair shaft elongation [12, 24].

Growth factors typically have a short half-life within hours to minutes, and strategies such as Fc-fusion engineering can extend serum half-life and sustain biological activity as long-acting growth factors [25]. The Fc-fusion approaches have shown favorable pharmacokinetics, pharmacodynamics, and efficacy in the context of human growth hormone therapeutics in reducing dosing frequency [25, 26].

Furthermore, hair care lotions containing extracts from *Centella asiatica* have been shown to strengthen hair roots [27]. Additional studies have indicated that *C. asiatica* extract promotes hair follicle health and enhances the inductive capacity of dermal papilla cells [28]. By increasing vascular endothelial growth factor (VEGF), *C. asiatica* improves the blood supply around the hair follicle, thereby supporting follicle health [29]. A purified compound from *C. asiatica*, araliadiol, has been found to promote cell proliferation and stimulate hair growth [30]. Plant-derived extracellular vesicle (EV), composed of nucleic acids, lipids, proteins, and secondary metabolites, contain many of the plant’s original bioactive ingredients and are readily absorbed by mammalian cells [31]. Therefore, we believe that EVs derived from *C. asiatica* represent an emerging natural agent capable of modulating local inflammation, activating dermal cells, and maintaining follicular stability for hair growth.

Caffeine, a commonly known alkaloid, has also been shown to stimulate the proliferation of hair matrix keratinocytes and counteract the inhibitory effects of dihydrotestosterone (DHT) [32, 33]. Caffeine promotes IGF-1 protein activity, effectively prolonging the anagen phase of the hair cycle, helping hair grow longer and stronger, and further supporting hair growth and follicle health [34]. By activating the intracellular cAMP signaling pathway, caffeine enhances follicular metabolism and stimulates growth potential. Additionally, its antioxidant properties help delay follicular degeneration, making it a promising ingredient for maintaining healthy hair growth [35]. Vitamin B5 (pantothenic acid) and its derivative D-panthenol have been implicated in hair growth regulation. In vitro studies demonstrated that D-panthenol promotes the proliferation of dermal papilla cells and outer root sheath cells, upregulating markers such as Ki-67, β-catenin, and VEGF, while suppressing TGF-β1 and caspase-associated apoptotic signals [36]. Additionally, animal studies have shown that pantothenic acid enhances hair follicle growth by upregulating ID3 and inhibiting the Notch pathway, accompanied by increased expression of IGF-1 and VEGF [37].

In this study, we developed a novel topical hair care serum that combines recombinant Fc-fusion long-acting insulin growth factor-1 (rIGF-1) and fibroblast growth factor-7 (rFGF-7), *C. asiatica* EV, with base formulation containing caffeine and panthenol, aiming to achieve a synergistic hair growth–promoting effect through multiple mechanisms of action. This formulation represents a unique integration of plant-derived biomaterial and recombinant long-acting growth factors, leveraging both hair invigorating signaling and targeted follicular stimulation. To our knowledge, this represented an unprecedented clinical study integrating plant EVs and long-acting growth factors in a human scalp care application. Here, we conducted a prospective, randomized, placebo-controlled human trial to evaluate the effects of this formulation in enhancing hair growth rate, density, and thickness, reducing hair loss, and improving scalp health.

## Materials and Methods

### Participants and Study Design

This study was a prospective, randomized, placebo-controlled, double-blinded clinical trial conducted to evaluate the effects of a topical scalp revitalizing essence on hair growth and hair loss. Participants were healthy adults of any gender aged 18 to 60 (inclusive) years.

Inclusion criteria include healthy adults defined as individuals without chronic diseases, significant illnesses (including cancer, post-stroke disorders, paralysis, acute myocardial infarction, coronary artery bypass surgery, end-stage renal disease, and major organ transplant or hematopoietic stem cell transplant), or allergic constitutions, and who are not currently taking any medication or using any scalp care products.

Exclusion criteria include: Individuals who are pregnant or breastfeeding, as well as males or females with chronic diseases, significant illnesses, or allergic constitutions, currently using scalp care products, students or employees affiliated with the principal investigator, individuals with scalp wounds, those who have participated in cosmetic product testing, and those who have undergone scalp aesthetic treatments within the past three months. Participants were randomly assigned to five groups (*n* = 12 per group) as detailed in the Description of Intervention below. This clinical study was registered at the ClinicalTrials.gov database as NCT06985121.

Participants were asked to apply approximately 1 mL of their assigned product once daily in the evening after shampooing. The product was applied over the entire scalp using a cotton pad and gently tapped into the scalp with fingertips to promote absorption. Participants were asked not to change their daily hair care routine other than the application of the test product. Usage was self-recorded in a provided diary. The duration of intervention was 8 weeks (56 days), with measurements taken at baseline (Day 0) and on Days 14, 28, 42, and 56.

### Description of Intervention

The following formulations were evaluated:

- Group A: Placebo control (Base formula without caffeine and panthenol, see below)
- Group B: Base formula (active ingredients: 0.1% caffeine and panthenol)
- Group C: Base formula + recombinant long-acting insulin growth factor-1 (rIGF-1) and recombinant long-acting fibroblast growth factor-7 (rFGF-7)
- Group D: Base formula + *Centella asiatica* extracellular vesicles (*C. asiatica* EV).
- Group E: Base formula + rIGF-1, rFGF-7, and *C. asiatica* EV. The composition of the base formula was as follows:

Aqua, glycerin, capryloyl glycine, xylitylglucoside, hexylene glycol, propanediol, caprylhydroxamic acid, 1,2-hexanediol, xanthan gum, menthol, menthyl lactate, PPG-10 methyl glucose ether, caffeine, and panthenol. Placebo control has an identical formulation to the base formula but without caffeine and panthenol, and is otherwise indistinguishable from other formulations. *C. asiatica* EVs were isolated as previously described with INCi names *Centella Asiatica* Leaf/Petiole Vesicles (INCI ID: 39425) and *Centella Asiatica* Callus Vesicles (INCI ID: 40060) [38]. rIGF-1 and rFGF-s7 are recombinant proteins modified for long-acting by fusing human IGF-1 or human FGF-7 with a human IgG1 Fc region fragment with a flexible linker with the following INCI names:

- rIGF-1: sr-(Methionyl sh Oligopeptide 2 Dipeptide-46 Oligopeptide-163 sh-Polypeptide 181), INCI ID:40196
- rFGF-7: sr-(Methionylsh-Polypeptide-3 Dipeptide-46 Oligopeptide-163 sh-Oligopeptide-197), INCI ID:40201

### Randomization and Blinding

Participants were randomly assigned in a 1:1:1:1:1 ratio into five parallel groups (n = 12 per group) using block randomization with a fixed block size of five, without stratification. The random allocation sequence was generated by the principal investigator and implemented using sequentially numbered, opaque, sealed envelopes (SNOSE) prepared by an independent staff member who was not involved in participant recruitment, assessment, or intervention delivery. The envelopes, identical in appearance and sealed with tamper-proof tape, were stored in a locked cabinet accessible only to the designated unblinded coordinator. Allocation was revealed only after eligibility confirmation and receipt of written informed consent.

A double-blind design was employed for this study. Participants, care providers administering the interventions, outcome assessors, and data analysts remained blinded to group allocation until the database lock, except in cases requiring emergency unblinding for safety reasons. Blinding was maintained by preparing all intervention and control products in identical containers with matched appearance, labeling, texture, viscosity, and scent. Each product was labeled only with a participant identification number, without indication of group assignment. The randomization code was securely held by the independent, unblinded coordinator and was not released until study completion.

### Assessment of Outcomes

Measurements were performed using two non-invasive diagnostic systems at standardized scalp sites (left, right, and vertex):

- Scalp sebum levels were quantified using C+K Multi Probe Adaptor MPA580 system with Sebumeter® SM815 probe (Courage-Khazaka Electronic GmbH, Koln, Germany) via grease-spot photometry.
- Hair length, thickness, and density were assessed with the ScalpX Intelligent Scalp Diagnostic System (VAST Technologies Inc., Taipei City, Taiwan) with a 5-megapixel digital microscope (DMC1213_USB 200X) to capture high-resolution scalp images and analyze using machine learning and artificial intelligence with an internal database.

Hair loss was assessment by standardized 60-stroke combing test [39]. Participants were instructed to wash their hair on the day prior to the assessment. On the following day, under dry hair conditions, the hair was combed 60 times using a standardized comb. The shed hairs were collected on a clean surface and subsequently counted manually.

All assessments were conducted onsite at Hungkuang University (Taichung City, Taiwan) under controlled environmental conditions (20 ± 1°C, 50 ± 5% RH) in a sealed room free of direct sunlight and airflow from vents.

For parameters such as sebum content, hair density, and hair loss, values were normalized as percentage improvement over baseline:

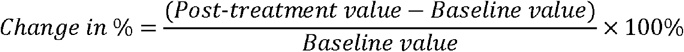

For hair length and hair thickness, absolute changes (in mm or μm) from baseline were used due to the linear nature of hair growth and narrow biological range, respectively.

Results for each participant were reported and calculated as the average of measurements from the left, right, and vertex regions of each participant to avoid pseudo-replication

### Statistical Analysis

Statistical analyses were performed using the built-in statistical package of GraphPad Prism 6.01 (Boston, MA, USA). Results were presented in mean values with 95% confidence intervals. For comparing demographic baseline values, χ2 test was used for gender and one-way ANOVA was used for continuous variables. To compare within-group changes over time, repeated measures ANOVA with Greenhouse-Geisser correction for sphericity followed by Dunnett’s post-hoc test for multiple comparisons, was used. For between-group comparisons at each timepoint, one-way ANOVA followed by Tukey’s HSD test for multiple comparisons. A p-value < 0.05 was considered statistically significant.

All tests were two-tailed. The sample size in this clinical study was not based on any statistical assumptions or calculations as this was an exploratory study aimed at observing the effects of the formulations.

## Results

### Participants

From 8 April 2025, to 31 July 2025, a total of 60 participants were enrolled and randomly assigned to five groups (*n* = 12 per group). All participants completed the study without major protocol violations or deviations. A CONSORT flow diagram summarizing participant recruitment, allocation, and follow-up is provided in Figure 1. The demographic characteristics and baseline values of the cohorts are shown in Table 1.

**Table 1.** Demographic characteristics and baseline values of the participants. *p-values calculated by χ2 test for gender and one-way ANOVA for baseline values.

| Allocated group | A | B | C | D | E | p-value* |
| --- | --- | --- | --- | --- | --- | --- |
| *n* | 12 | 12 | 12 | 12 | 12 | ns |
| Gender |  |  |  |  |  | ns |
| Female | 11 (91.7%) | 10 (83.3%) | 10 (83.3%) | 10 (83.3%) | 10 (83.3%) |  |
| Male | 1 (8.3%) | 2 (16.7%) | 2 (16.7%) | 2 (16.7%) | 2 (16.7%) |  |
| Baseline values (mean ± SD) | | | | | | |
| Age (years) | 36.3 ± 8.5 | 34.7 ± 10.5 | 38.3 ± 10.3 | 36.2 ± 11.1 | 36.3 ± 8.7 | ns |
| Sebum content (a.u.) | 58.0 (24.6) | 49.9 (20.2) | 62.8 (23.3) | 56.2 (13.5) | 47.8 (11.0) | ns |
| Hair length (cm) | 36.6 (17.3) | 33.0 (14.6) | 35.1 (16.8) | 38.5 (14.3) | 33.0 (15.1) | ns |
| Hair thickness (μm) | 75.8 (6.3) | 73.4 (8.5) | 70.3 (9.8) | 76.3 (5.8) | 71.0 (5.9) | ns |
| Hair density (no. of hairs/cm<sup>2</sup>) | 146.1 (11.3) | 148.5 (16.9) | 133.7 (14.5) | 146.6 (21.7) | 153.5 (14.2) | ns |
| Hair loss (no. of hairs lost/assessment) | 13.3 (5.4) | 15.6 (5.6) | 15.0 (4.1) | 16.4 (6.2) | 16.1 (5.1) | ns |

**Figure 1.**
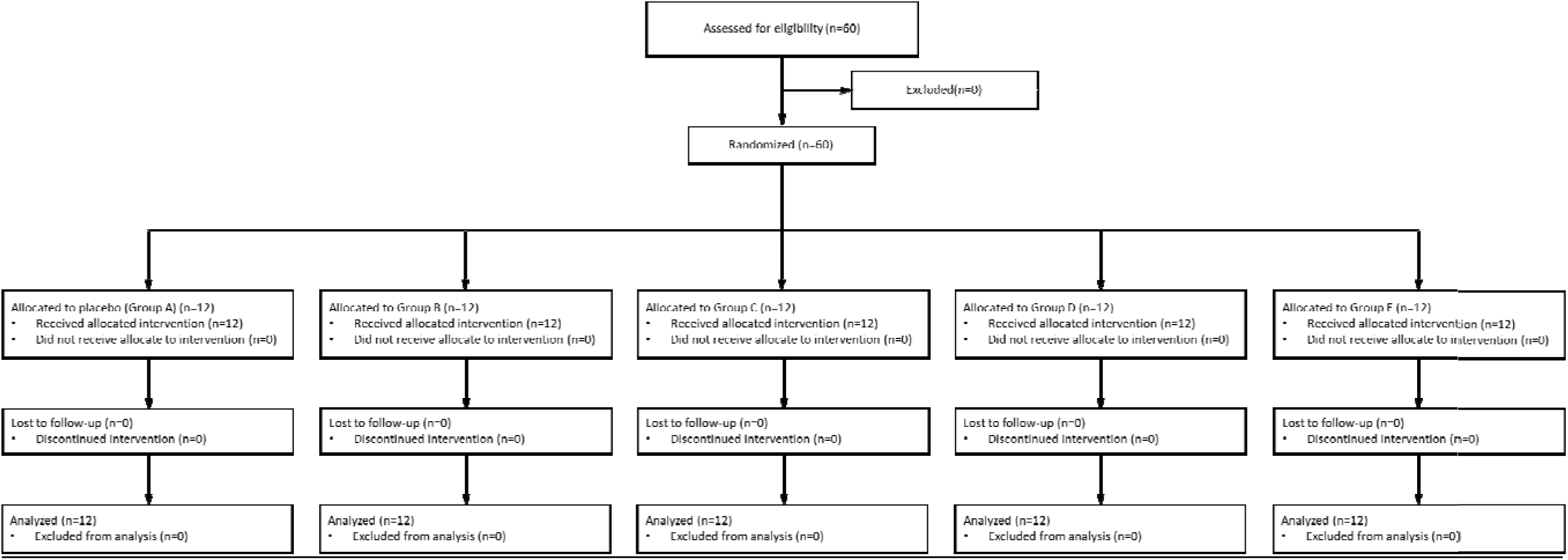
CONSORT flowchart of the study.

### Assessments of Outcomes

#### Sebum Content

Sebum levels were found to decrease over time in all groups, with formulations containing *C. asiatica* EV with rIFG-1 and rFGF-7 (Group E) resulting in significantly lower levels of sebum than the base formulation (Group B) and placebo control by Day 56 (p < 0.05 and p < 0.01, respectively) (Figure 2). By Day 56, Group E showed an average reduction of 59% in sebum from baseline, compared to placebo (47.4% reduction). In contrast, minimal change was observed in Groups B to D compared to placebo (results statistically not significant). Thus, the combined use of *C. asiatica* EVs, IGF-1, and FGF-7s could reduce sebum production in the scalp as observed.

**Figure 2.**
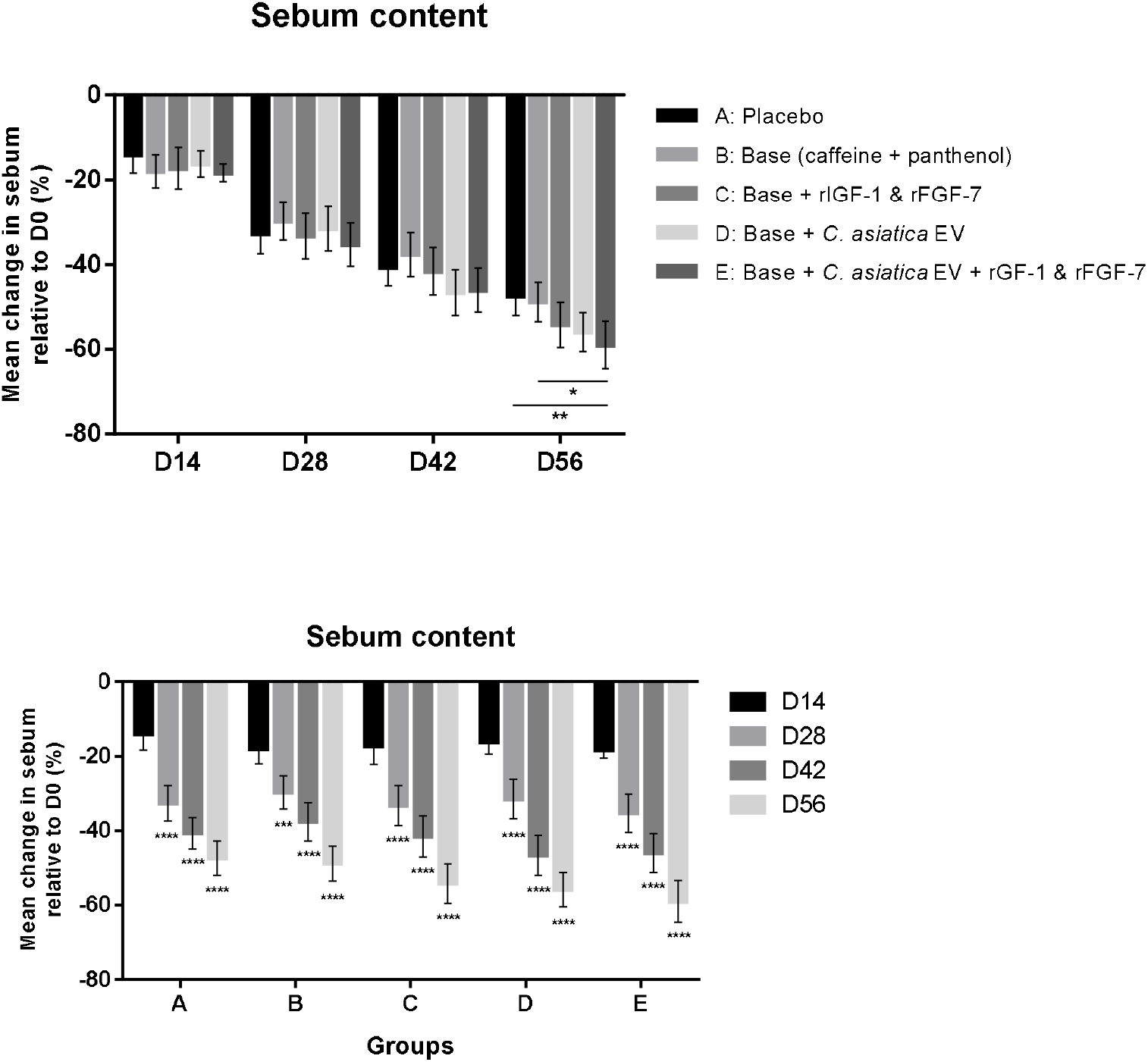
Change (%) in sebum content compared to baseline (Day 0) at 14, 28, 42, and 56 days after start of the study. Top: Comparison of between-group at each timepoint with statistical significance calculated using one-way ANOVA followed by Tukey’s HSD test; Bottom: Comparison of within-group changes over time with statistical significance calculated using repeated measures ANOVA followed by Dunnett’s post-hoc test. * p < 0.05, ** p < 0.01, **** p < 0.0001.

### Hair Length

As expected, an increase in hair length was observed in all groups, reflecting natural hair growth (Figure 3 bottom). As shown in Figure 3, top, Group E achieved the highest mean cumulative growth by Day 56 (3.5 cm), significantly exceeding both placebo and Group B (p < 0.001 and p < 0.01, respectively). Interestingly, the differences in hair length growth were already significant between Group E and placebo (p < 0.0001) on Day 14, and continued to grow at a faster pace than placebo control at all other time points throughout the study (Figure 3). These results suggest an acceleration of hair growth in groups receiving growth factors and/or EVs.

**Figure 3.**
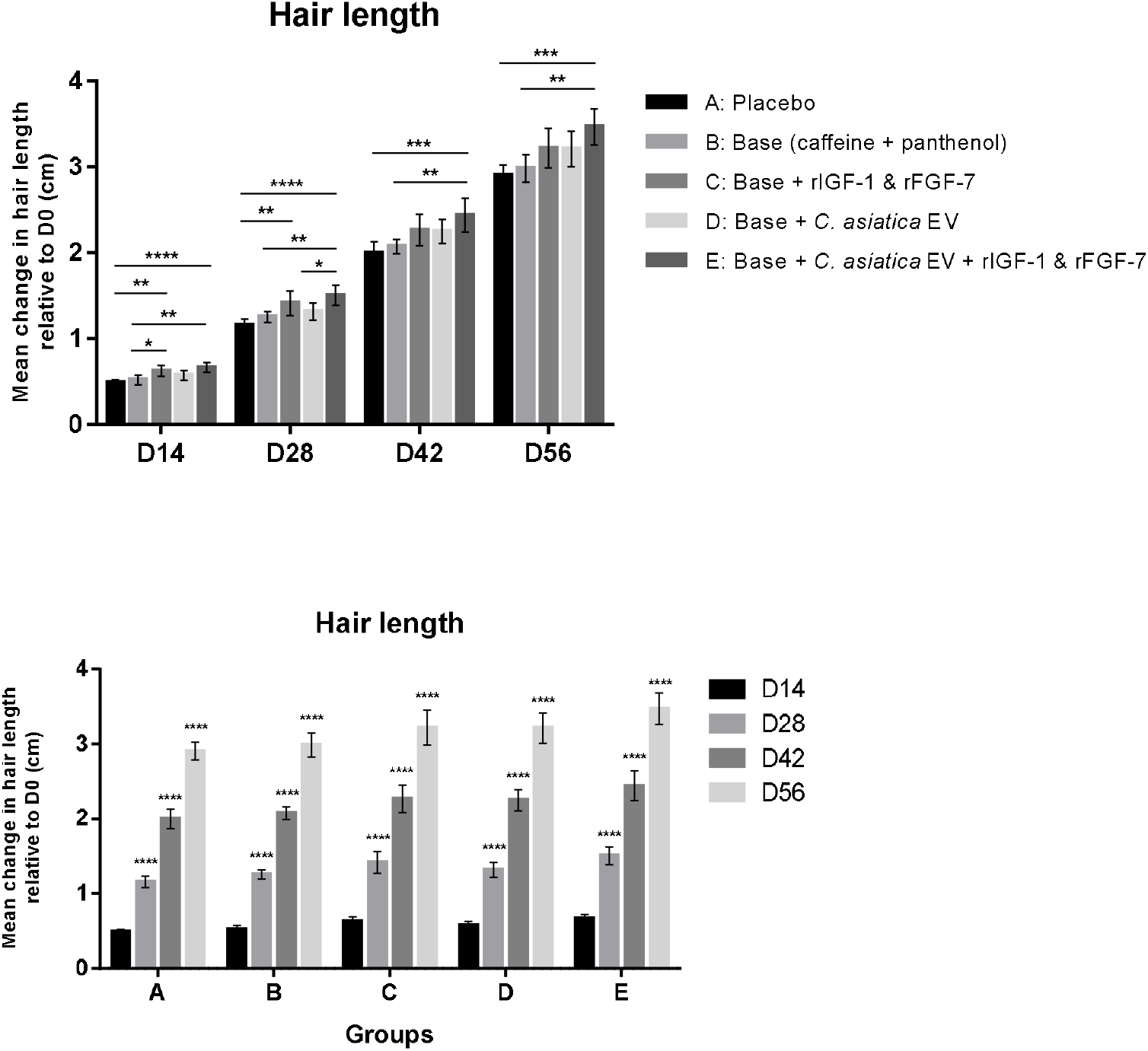
Change (cm) in hair length compared to baseline (Day 0) at 14, 28, 42, and 56 days after start of the study. Top: Comparison of between-group at each timepoint with statistical significance calculated using one-way ANOVA followed by Tukey’s HSD test; Bottom: Comparison of within-group changes over time with statistical significance calculated using repeated measures ANOVA followed by Dunnett’s post-hoc test. * p < 0.05, ** p < 0.01, *** p < 0.001, **** p < 0.0001.

### Hair Thickness

All treatment groups showed significant increases in hair shaft diameter over the study period (Figure 4, bottom). Group E again exhibited the greatest effect, with a mean increase of 27.9 μm at Day 56 compared to 13.9 μm of placebo (p < 0.0001) (Figure 4 top). The other formulations also showed marked thickening of hair shafts compared to placebo (p < 0.05). Noticeably, the addition of rIGF-1 and rFGF-7 to *C. asiatica* EV in Group E resulted in a significant increase (p < 0.05) in hair thickening compared to *C. asiatica* EV only in Group D (27.9 μm vs 19.3 μm).

**Figure 4.**
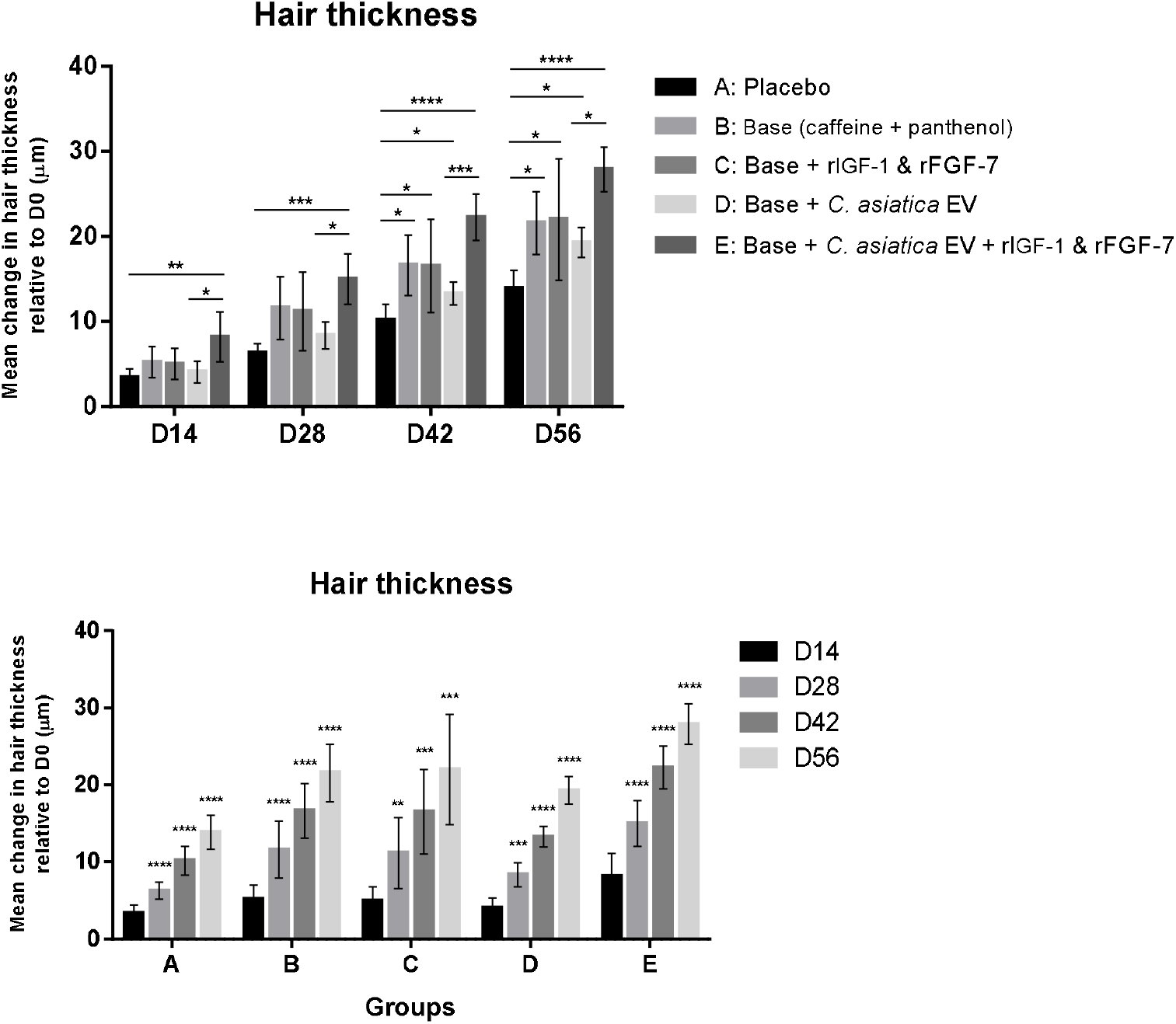
Change (µm) in hair thickness compared to baseline (Day 0) at 14, 28, 42, and 56 days after start of the study. Top: Comparison of between-group at each timepoint with statistical significance calculated using one-way ANOVA followed by Tukey’s HSD test; Bottom: Comparison of within-group changes over time with statistical significance calculated using repeated measures ANOVA followed by Dunnett’s post-hoc test. * p < 0.05, ** p < 0.01, *** p < 0.001, **** p < 0.0001.

### Hair Density

Hair density increased over time in all treatment groups, including the placebo group (Figure 5, bottom). Group E showed the most pronounced improvement and, with a mean increase of approximately double (23.9% vs 11.9% increase relative to baseline, p < 0.001) than that of placebo by Day 56 (Figure 5 top). Group C also showed significant increases compared to placebo (23.2% vs 11.9%, p < 0.01), while Groups B and D showed moderate but significant improvement (20.3% vs 11.9%; 20.0% vs 11.9%, p < 0.05).

**Figure 5.**
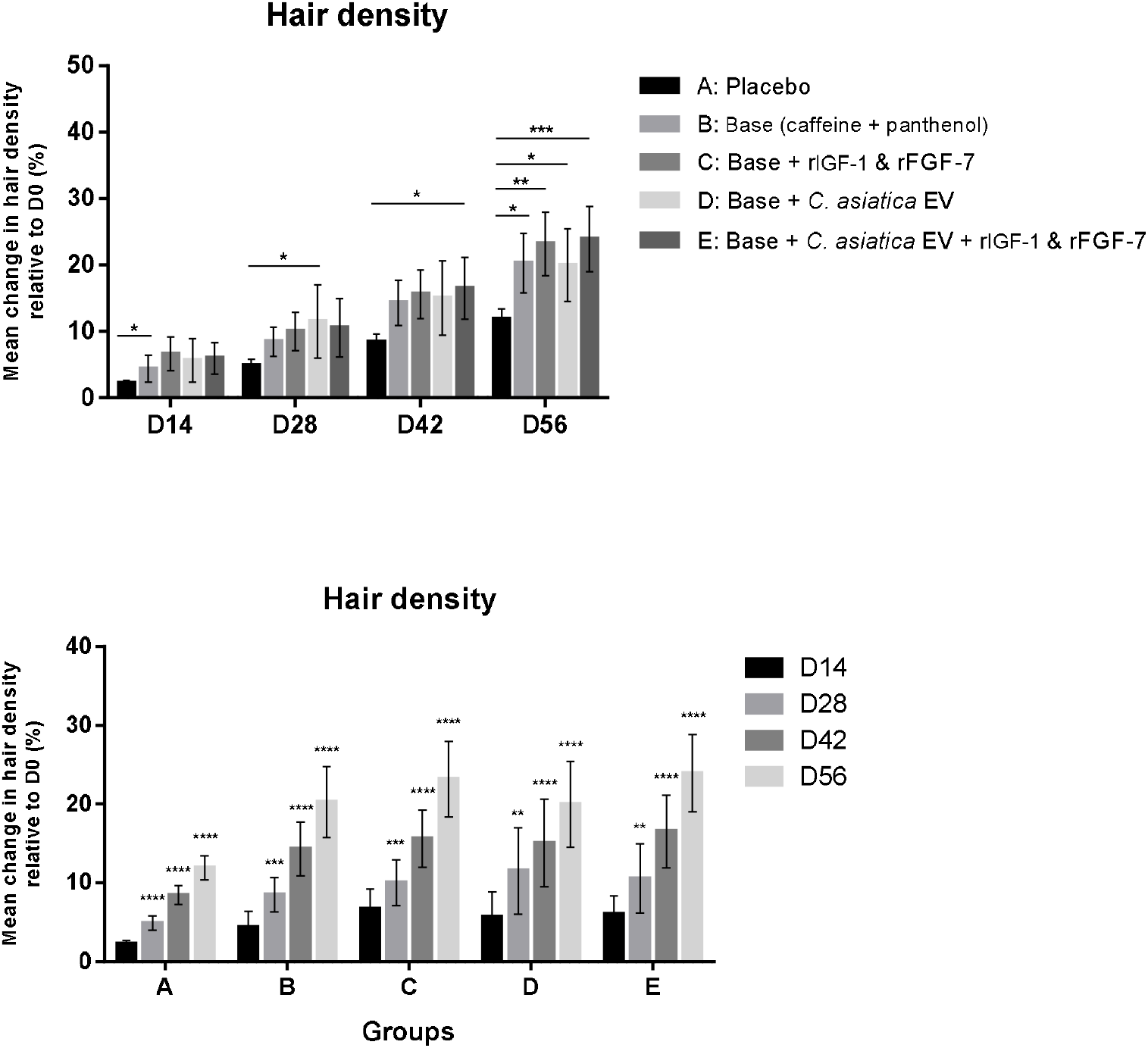
Change (%) in hair density compared to baseline (Day 0) at 14, 28, 42, and 56 days after start of the study. Top: Comparison of between-group at each timepoint with statistical significance calculated using one-way ANOVA followed by Tukey’s HSD test; Bottom: Comparison of within-group changes over time with statistical significance calculated using repeated measures ANOVA followed by Dunnett’s post-hoc test. * p < 0.05, ** p < 0.01, *** p < 0.001, **** p < 0.0001.

### Hair Loss

The percentage of hair loss decreased in all treatment groups during the course of the study, even in the placebo group (Figure 6 bottom). Group E had the best reduction in hair loss on Days 42 (53.6% vs 32.5%) and 56 (63.6% vs 43.1%) compared to placebo (p < 0.01 and p < 0.05, respectively) (Figure 6 top). Other formulations did not result in significantly decreased hair loss compared to placebo at any point in time during the study.

**Figure 6.**
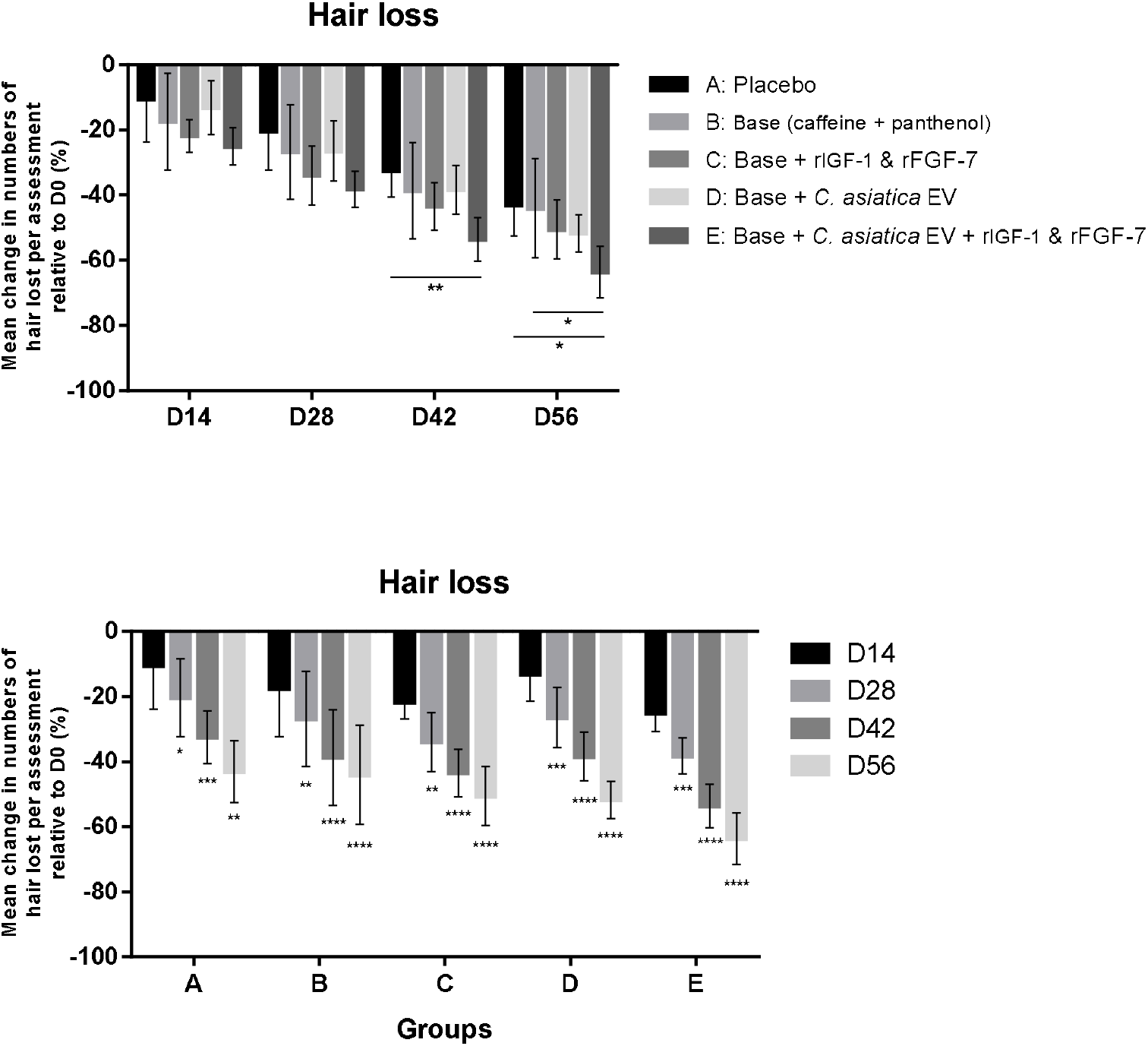
Change (%) in hair lost during the assessment event compared to baseline (Day 0) at 14, 28, 42, and 56 days after start of the study. Top: Comparison of between-group at each timepoint with statistical significance calculated using one-way ANOVA followed by Tukey’s HSD test; Bottom: Comparison of within-group changes over time with statistical significance calculated using repeated measures ANOVA followed by Dunnett’s post-hoc test. * p < 0.05, ** p < 0.01, *** p < 0.001, **** p < 0.0001.

Figure 7 above shows the before-and-after photographs of the participants in Group E at the beginning (Day 0) and after 56 days of treatment, which resulted in visible increase in overall hair density, especially at the vertex.

**Figure 7.**
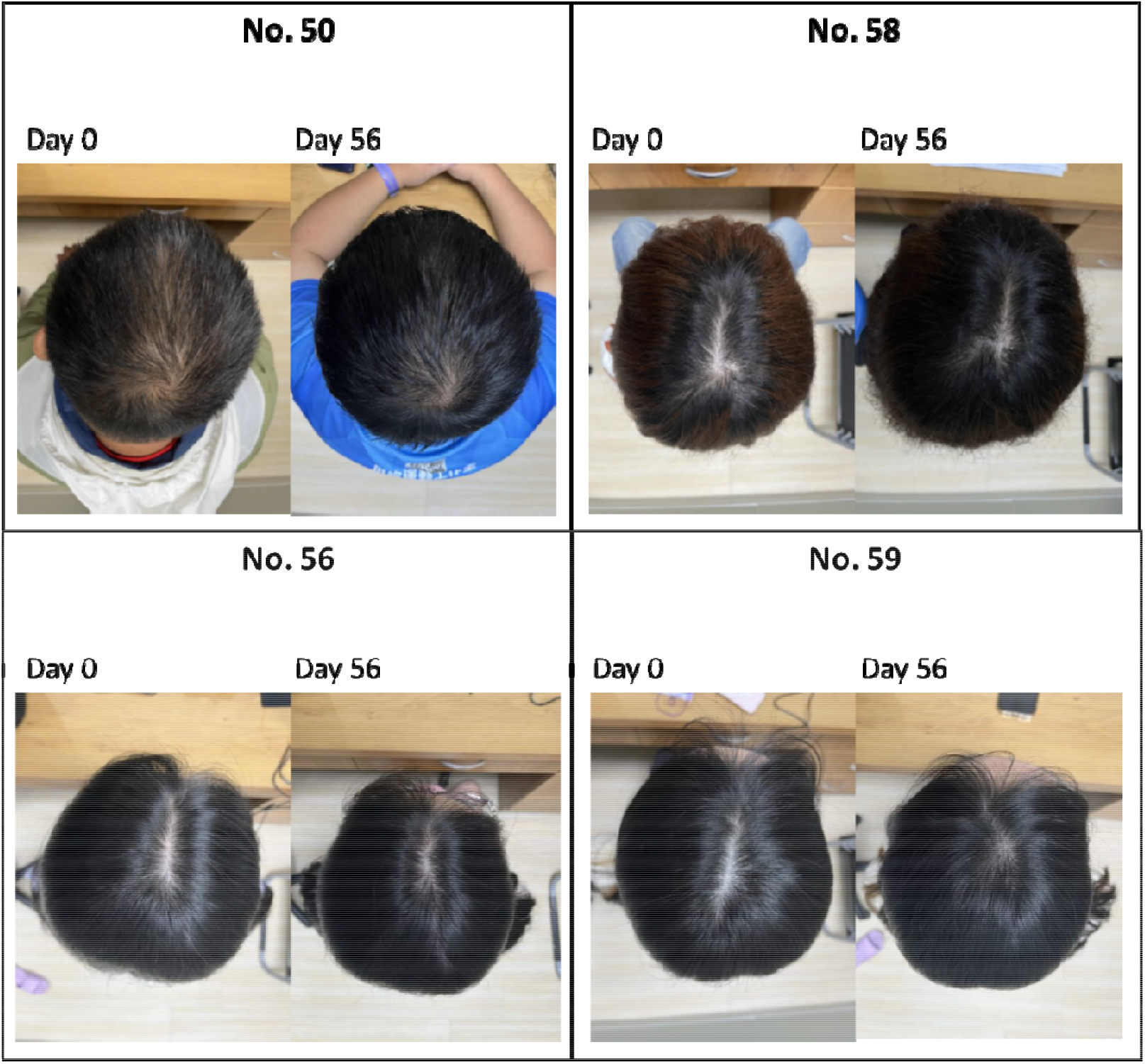
Representative photographs of the participants in Group E (Base formula with rIGF-1, rFGF-7, and *C. asiatica* EV) on Day 0 and Day 56 after treatment.

## Discussion

This randomized placebo-controlled clinical trial evaluated the efficacy of formulations of scalp revitalizing essence containing rIGF-1, rFGF-7, *C. asiatica* EVs, caffeine, and panthenol in improving scalp and hair parameters over a 56-day period. Our findings demonstrate that the formulation resulted in measurable improvements, with Group E (the full formulation) showing the most consistent and pronounced effects in sebum reduction, hair growth (length, thickness, and density) and hair loss reduction. For example, by Day 56, hair shaft thickness increased by 27.9 μm, significantly outperforming placebo (13.9 μm, p < 0.0001) and other formulations (p < 0.05). Similarly, hair density increased by 23.9%, nearly double that of the placebo group (p < 0.001). These findings are in line with our proposed synergistic action of the combined active ingredients of *C. asiatica* EV, rIGF-1, and rFGF-7, which are known to modulate signaling pathways involved in follicle development and dermal papilla function [11–15, 27-29].

Hair length results further supported this, with Group E showing a cumulative growth of 3.5 cm by Day 56, significantly exceeding placebo (p < 0.001) and showing differences as early as Day 14 (p < 0.0001) (Figure 3). While hair naturally grows at a relatively fixed rate, the enhancement observed here may be attributable to follicular stimulation via growth factor signaling pathways by IGF-1 and FGF-7 [13-14, 17].

In terms of hair loss, Group E demonstrated a 63.6% reduction by Day 56 compared to 43.1% in placebo (p < 0.05). Interestingly, while all groups experienced some degree of hair loss reduction, only Group E achieved statistically significant improvements over placebo. This is consistent with IGF-1’s known role in inhibiting follicular apoptosis [18, 19] and caffeine’s ability to counteract DHT-mediated suppression of hair matrix keratinocyte activity [32, 33].

Sebum production was reduced in all groups, but only significantly in Group E with 59% reduction relative to baseline, significantly outperforming both placebo (p < 0.01) and the base formulation Group B (p < 0.05). *C. asiatica* EVs are known to be rich in bioactive lipids, proteins, and anti-inflammatory compounds as well as novel mRNAs, which may contribute to homeostatic regulation of sebaceous glands and microvascular perfusion [29, 31, 40]. While the effect of *C. asiatica* EVs on hair follicles has yet to be reported, *C. asiatica* extract is known to promote dermal papilla activity and vascular growth via VEGF signaling [28, 29].

Our results obtained here are in agreement with previous studies demonstrating the roles of IGF-1 and FGF-7 in promoting dermal papilla proliferation, follicle regeneration, and keratinocyte survival [11–14, 16–18]. While topical minoxidil and finasteride remain the most widely used growth stimulants, they are known to cause rebound shedding upon discontinuation and side effects that impact quality of life [6-8]. Therefore, combining growth factor with plant-derived EVs may offer a multi-targeted approach to scalp care. To our knowledge, this is the first human trial to evaluate a combined formulation of plant-derived EVs and growth factors that explores the regenerative potential of bio-signaling molecules in a topical, non-pharmaceutical format for cosmetic application [4, 5, 9, 11].

Here we discuss several limitations of the study. First, a modest improvement in hair density and hair loss was observed in the placebo group even in the absence of active ingredients such as IGF-1, FGF-7, *C. asiatica* EVs, likely attributable to natural telogen-to-anagen cycling, standardized hair care routines, and psychological effects of study participation [2-3, 41]. Some components within the placebo composition may also contribute to the placebo effect, even though they are not active ingredients *per se*. Glycerin is a commonly used humectant that can retain water in the stratum corneum and help skin barrier repair, leading to improved scalp hydration [42]. Menthol and menthyl lactate can activate TRPM8 channels to provide a cooling sensation while also stimulating cutaneous blood flow, possibly leading to increased perfusion of follicular metabolism and reduced skin irritation [43]. Second, the short duration (56 days) of the study was another limitation, which may not fully capture long-term follicular cycling or delayed responses. Third, the relatively young age of participants (mean age 36 years) and heavily skewed toward female may limit the generalizability of the findings to older populations with more advanced androgenetic alopecia or age-related scalp conditions [1, 4]. Since follicular renewal capacity declines with age, further studies in middle-aged and older individuals using a balanced gender ratio are warranted to confirm whether the synergistic effects of *C. asiatica* EVs, rIGF-1 and rFGF-7 are equally effective across different age groups. Additionally, the small sample size precludes subgroup comparisons or regression analysis by age or scalp condition. Future studies should explore long-term safety and outcomes, mechanistic biomarkers, and scalp biopsies to further support the long-term and increased frequency of use of the product.

## Conclusions

The topical application of an essence containing *C. asiatica* EVs, IGF-1, FGF-7, and caffeine led to significant improvements in hair thickness, density, length, and sebum control, along with a reduction in hair loss after 56 days of use. These effects suggest a synergistic interaction between plant-derived EVs, rIGF-1, and rFGF-7, offering a novel, practical, and effective strategy for maintaining routine scalp and hair health.

## Author Contributions

Conceptualization, supervision and project administration: T.-M.C., C.C., T.-Y.K. Visualization and formal analysis: T.-M.C., W.-J. H., L.T.-C.L. Investigation, data acquisition and curation: T.-M.C., H.-C.H., J.-Y. L. Methodology: T.-M.C., C.-C.W., H.-C.H., J.-Y. L., C.-H.C., P.-L.K., W.-H.T., W.-Y.Q. I.P., T.-Y.K.

Resources: C.-C.W., C.-H.C., P.-L.K., W.-H.T., T.-Y.K. Writing—original draft preparation: T.-M.C., W.-J. H., L.T.-C.L. Writing—review and editing: T.-M.C., W.-J. H., L.T.-C.L., W.-Y.Q., I.P., C.C., T.-Y.K. All authors reviewed the manuscript.

## Funding

Schweitzer Biotech Company provided the necessary funding for the study as well as provided the test products, but had no role in enrollment, participation, data collection, data analysis, or completion of final report of the clinical study.

## Institutional Review Board Statement

The finalized protocol, case report form, advertisement, and informed consent form were reviewed and approved by the institutional review board of the Antai Tian-Sheng Memorial Hospital (IRB No. 25-016-A, approved February 06, 2025). All participants provided written informed consent prior to the start of the study. This study was conducted in compliance with Declaration of Helsinki and in compliance with all International Conference on Harmonization Good Clinical Practice Guidelines.

## Informed Consent Statement

All participants provided written informed consent prior to the start of the study.

## Data Availability

All relevant data is provided within the manuscript

## Acknowledgements

We thank colleagues at Schweitzer Biotech Company for reviewing and providing feedback for the manuscript preparation.

## Conflicts of Interest

C.-C. W., C.-H. C., P.-L. K., W.-H. T., W.-J. H., L. T.-C. L., C. C., and T.-Y. K. are employees of Schweitzer Biotech Company. W.-Y. Q. and I. P. are consultants employed by Schweitzer Biotech Company. T.-M. C. and J.-Y. L. have no competing interests to declare.

## Notes

### Clinical Trial

NCT06985121

### Author Declarations

Institutional Review Board of the Antai Tian-Sheng Memorial Hospital

